# Global longitudinal strain and plasma biomarkers for prognosis in heart failure complicated by diabetes

**DOI:** 10.1101/2023.03.20.23287472

**Authors:** Nithin R. Iyer, Siew-Pang Chan, Oi Wah Liew, Jenny P.C. Chong, Jennifer A. Bryant, Thu-Thao Le, Chanchal Chandramouli, Patrick J. Cozzone, Frank Eisenhaber, Roger Foo, A. Mark Richards, Carolyn S.P. Lam, Martin Ugander, Calvin W-L. Chin, the ATTRaCT investigators

**Author notes:** **Address for Correspondence**, Calvin W-L. Chin, MD PhD, Department of Cardiology, National Heart Centre Singapore, 5 Hospital Drive Singapore 169609, E. denotes equal contribution (Martin Ugander, Calvin W-L. Chin).

## Abstract

**Background:** Heart failure (HF) and diabetes are associated with increased incidence and worse prognosis of each other. The prognostic value of global longitudinal strain (GLS) measured by cardiovascular magnetic resonance (CMR) has not been established in HF patients with diabetes.

**Methods:** Consecutive patients (n=315) with HF underwent CMR at 3T, including GLS, late gadolinium enhancement (LGE), native T1, and extracellular volume fraction (ECV) mapping. Plasma biomarker concentrations were measured including: N-terminal pro B-type natriuretic peptide(NT-proBNP), high-sensitivity troponin T(hs-TnT), growth differentiation factor 15(GDF-15), soluble ST2(sST2) and galectin 3(Gal-3). The primary outcome was a composite of all-cause mortality or HF hospitalisation.

**Results:** Compared to those without diabetes (n=156), the diabetes group (n=159) had a higher LGE prevalence (76 vs 60%, p<0.05), higher T1 (1285±42 vs 1269±42ms, p<0.001) and higher ECV (30.5±3.5 vs 28.8±4.1%, p<0.001). The diabetes group had higher NT-pro-BNP, hs-TnT, GDF-15, sST2 and Gal-3. Diabetes conferred worse prognosis (hazard ratio (HR) 2.33 [95% confidence interval (CI) 1.43-3.79], p<0.001). In multivariable Cox regression analysis including clinical markers and plasma biomarkers, sST2 alone remained independently associated with the primary outcome (HR per 1 ng/mL 1.04 [95% CI 1.02-1.07], p=0.001). In multivariable Cox regression models in the diabetes group, both GLS and sST2 remained prognostic (GLS: HR 1.12 [95% CI 1.03-1.21], p=0.01; sST2: HR per 1 ng/mL 1.03 [95% CI 1.00-1.06], p=0.02).

**Conclusions:** Compared to HF patients without diabetes, those with diabetes have worse plasma and CMR markers of fibrosis and a more adverse prognosis. GLS is a powerful and independent prognostic marker in HF patients with diabetes.

## Background

The Asian continent has the highest prevalence of heart failure (HF) cases globally [1]. Diabetes is especially common in South East Asian HF populations, where a unique ‘lean-diabetic’ phenotype with worse outcomes has been identified [2]. The Asian Sudden Cardiac Death in Heart Failure (ASIAN-HF) registry (across 11 Asian countries) reported a 42.5% prevalence of diabetes among HF patients, specifically in higher-income countries such as Singapore, Hong Kong, and Thailand [3].

The diabetic heart is characterised by a number of structural abnormalities including diffuse interstitial myocardial fibrosis, myocyte hypertrophy, and impaired coronary microvascular perfusion, which have all been implicated in the development of both diastolic and systolic dysfunction [4, 5]. Patients with HF and diabetes have consistently worse clinical outcomes, including higher risk of hospitalisation for HF and death, compared to those without diabetes [6–8]. These findings appear to hold regardless of whether the HF is ischaemic or non-ischaemic in etiology, and regardless of left ventricular (LV) ejection fraction [7, 8].

Cardiac magnetic resonance (CMR) imaging has become the non-invasive reference standard for evaluating HF due to its ability to accurately assess cardiac morphology, function, and myocardial tissue characteristics. In particular, late gadolinium enhancement (LGE) permits visualisation of focal replacement myocardial fibrosis, while T1 mapping pre- and post-gadolinium contrast enables non-invasive measurement of the myocardial extracellular volume fraction (ECV), a quantitative marker of myocardial diffuse interstitial fibrosis. Additionally, global longitudinal strain (GLS), defined as the change in the LV myocardial length between diastole and systole divided by the original end-diastolic length, provides a measure of LV systolic function by CMR that is effectively the same as GLS measured by echocardiography [9, 10].

Patients with diabetes frequently have impaired GLS and have a higher degree of myocardial fibrosis as assessed by ECV and histology [11, 12]. GLS by CMR appears to have prognostic value in HF regardless of ejection fraction and whether the cause of HF is ischemic or non-ischemic [13–15]. There is a growing body of literature demonstrating the prognostic value of GLS in asymptomatic patients with diabetes [16, 17]. However, the prognostic utility of GLS in diabetes patients with established HF failure is unknown.

Growth differentiation factor-15 (GDF-15), soluble ST2 (sST2) and galectin 3 (Gal-3) are circulating plasma biomarkers associated with inflammation, fibrosis and cardiac remodelling [18]. Plasma concentrations of these biomarkers appear to provide prognostic information in HF patients beyond established markers including cardiac troponins and natriuretic peptides [18–21]. The prognostic relevance of these biomarkers has not yet been established in HF patients with diabetes.

This study aimed to assess the prognostic significance of GLS by CMR and novel HF plasma biomarkers associated with inflammation and fibrosis in a cohort of patients with HF and diabetes across the spectrum of LV ejection fraction. We hypothesized that GLS would have an incremental prognostic association in this group, beyond plasma HF biomarkers, LV ejection fraction and CMR markers of myocardial fibrosis.

## Methods

### Study Population

Patients with HF were recruited prospectively across six tertiary cardiac centres in Singapore (Asian neTwork for Translational Research and Cardiovascular Trials [ATTRaCT], ClinicalTrials.gov NCT02791009). Patients were included if they presented to hospital with a primary diagnosis of HF, or if they attended a hospital clinic within 6 months of an episode of decompensated HF (requiring hospitalization or treatment in an out-patient setting). In all cases, a trained cardiologist adjudicated the clinical diagnosis of HF. The exclusion criteria were: HF primarily due to severe valve disease, HF due to acute coronary syndrome resulting in a transient episode of acute pulmonary oedema, severe renal failure (estimated glomerular filtration rate < 15 mL/min per 1.73m^2^), specific causes of HF (constrictive pericarditis, complex adult congenital heart disease, hypertrophic cardiomyopathy, eosinophilic myocarditis, cardiac amyloidosis, and acute chemotherapy-induced cardiomyopathy), isolated right HF, and life threatening non-cardiac co-morbidity with life expectancy of <1 year. All patients underwent clinical assessment at baseline. Diabetes status was identified by baseline questionnaire at recruitment.

### Biomarkers

Blood was collected in dipotassium (K_2_)-EDTA vacutainer tubes and transported on ice for processing within one hour. Plasma was separated by centrifugation at 3500g for 10 minutes at 4°C and stored at -80°C until analysis.

Plasma N-terminal pro B-type natriuretic peptide (NT-proBNP) and high-sensitivity troponin T (hs-TnT) were measured by electrochemiluminescence immunoassay using the Elecsys proBNP G2 V2.1 and Elecsys Troponin T hs V2.1 assays on the Cobas e411 immuno-analyser (Roche Diagnostic GmbH, Mannheim, Germany). The measurement ranges of NT-proBNP and hs-TnT were 10-35000 pg/ml and 3-10000 pg/ml, respectively. Laboratory average concentration and inter-assay coefficient of variation (%CV) of low (NT-proBNP: 147 pg/ml, 4.48%; hs-TnT: 26.8 pg/ml, 5.05%) and high (NT-proBNP: 4679 pg/ml, 4.97%; hs-TnT: 2120 pg/ml, 3.69%) quality control samples of the NT-proBNP and hs-TnT assays were established over 84 and 73 independent assays, respectively.

Human GDF-15 (R&D Systems, Minneapolis, MN, USA; Cat#DGD150), sST2 (Presage ST2 assay, Critical Diagnostics, California, USA) and Gal-3 (R&D Systems, Minneapolis, MN, USA; Cat#DGAL30) were measured by ELISA on the Enspire Multimode Microplate Reader (Perkin Elmer, Waltham, MA, USA). Results were interpolated from standard curves fitted on 5-parameter logistic model (5-PL) using the instrument’s Enspire®software. Laboratory inter-assay %CV of quality control samples were 7.41% at 126 pg/ml, 7.71 % at 360 pg/ml and 8.43% at 778 pg/ml for GDF-15 (n=98), 18.0% at 30.0 ng/ml and 15.2% at 63.6 ng/ml for sST2 (n=98) and 10.4% at 0.83 ng/ml, 10.6% at 2.41 ng/ml and 12.3% at 4.82 ng/ml for Galectin-3 (n=56).

### Clinical Outcomes

The primary outcome was a composite of time to either first hospitalization for HF (regardless of prior history of hospitalization for HF) or all-cause mortality. Follow-up was conducted through a clinic visit at 6 months after baseline assessment and structured phone interviews with the participants at the 1- and 2-year timepoints. Data in patients who were lost to follow-up were censored at the date when the patient was last known to be alive and had not experienced an event.

### Cardiovascular magnetic resonance image acquisition

All patients in the ATTRaCT cohort were invited and assessed for suitability for CMR. Those who agreed and were eligible underwent a standardized CMR protocol with a 3 Tesla MRI scanner (Ingenia, Philips Healthcare, Best, The Netherlands). Balanced steady-state free precision cines were acquired in the standard long-axis views and a short-axis stack from base to apex, as described previously [22]. LGE images were acquired at 10 min after 0.1 mmol/kg of gadobutrol (Gadovist®, Bayer Pharma AG, Germany) with a phase sensitive inversion-recovery gradient-echo imaging sequence. Typical parameters were: repetition time (TR) = 6.1 ms; echo time (TE) = 3 ms; time to inversion (TI) = 320–340 ms, flip angle 25°, voxel size = 1.5×1.7×8 mm, SENSE factor = 2.4, slice thickness 8mm with 2mm gap to match short-axis cine slice positions. The inversion time for optimal myocardial nulling was selected from an inversion time scout sequence. T1 maps were acquired at the basal and mid-ventricular short-axis levels, pre- and 15-min post-contrast with modified Look-Locker Inversion-recovery (MOLLI) 5s(3s)3s and 4s(1s)3s(1s)2s acquisition schemes, respectively [23].

### CMR analysis

Image analysis was performed using CVI42 software (Circle Cardiovascular Imaging, Calgary, Canada) by trained imaging fellows at the National Heart Research Institute of Singapore CMR Core Laboratory, who were blinded to the clinical information of the patients. Ventricular volumes, mass and ejection fraction were measured from the short-axis cine stack, using manual contouring of the left ventricle in end-diastole and end-systole, excluding papillary muscles, as detailed previously [22]. LV volumes and mass data were indexed to body surface area. The presence of LGE was assessed qualitatively by two readers according to the recommendations by the Society of CMR [24]. Average native and post-contrast myocardial T1 values were measured by placing a region of interest (ROI) within the middle third of the short-axis myocardial wall at the basal- and mid-ventricular levels, while avoiding regions of focal LGE. The myocardium-blood pool interface was carefully avoided in order to prevent partial volume effects. Pre- and post-contrast blood T1 values were measured in a ROI drawn within the blood pool. ECV was calculated from the pre- and post-contrast average blood and myocardial T1 values, as described previously [25, 26]. Myocardial strain was analysed in the cine images using the Tissue Tracking Plugin [27].

### Statistics

Normality was assessed for continuous variables using the Shapiro-Wilk test. Normally distributed data are presented as mean ± standard deviation. Non-normally distributed data are presented as median [interquartile range]. Comparisons were performed for continuous variables using the parametric Student *t*-test or the non-parametric Mann-Whitney U test. Categorical variables are presented as number (percentage) and compared using the χ^2^ test.

Univariable Cox regression analysis was performed to identify prognostic variables in the entire cohort and also in the diabetes subgroup. Clinically relevant variables (age, sex, body mass index, diabetes, smoking, coronary artery disease, hypertension, New York Heart Association functional class and systolic blood pressure), circulating biomarkers (NT-proBNP, hs-TnT, GDF-15, sST2 and Gal-3) and CMR markers of function, remodelling and fibrosis (LV end diastolic volume index, mass index, ejection fraction, GLS, LGE, Native T1, ECV) were tested in the Cox models. Covariates with a p-value <0.05 in univariable analyses were entered into the multivariable Cox model to identify independently prognostic variables, using forward stepwise selection. Event-free survival curves were examined using the Kaplan-Meier method and compared with the log-rank test. Statistical analyses were performed using SPSS Version 28 (Statistical Package for the Social Sciences, International Business Machines, Inc., Armonk, New York, USA) and GraphPad Prism 9.4.1 (GraphPad Software, Inc., San Diego, California, USA). A two-sided p-value <0.05 was considered as statistically significant.

## Results

Figure 1 shows a flow chart describing patient inclusion. Of the 623 patients enrolled in the ATTRaCT study, 523 patients underwent a baseline CMR study. We excluded 168 studies performed without contrast. An additional 40 patients were excluded due to incomplete baseline clinical or CMR data sets, alternative diagnosis on the basis of CMR, or loss to follow-up. In total, 315 subjects (diabetes, n=159; without diabetes, n= 156) were included in the study cohort.

**Figure 1.**
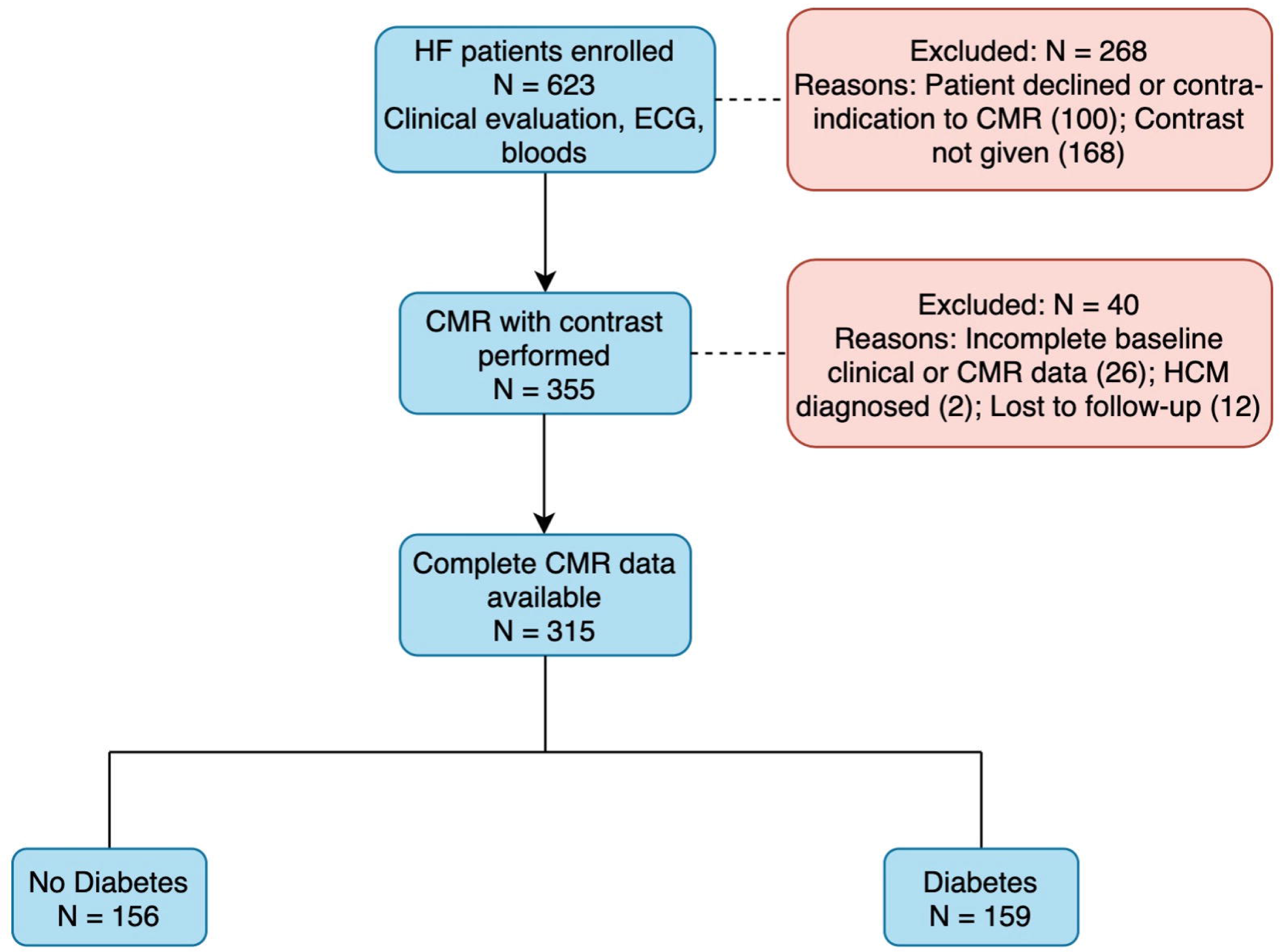
Flow chart of patient inclusion. Abbreviations: HF: heart failure; ECG: 12-lead electrocardiogram; CMR: Cardiovascular magnetic resonance; HCM: Hypertrophic cardiomyopathy.

The baseline clinical characteristics are shown in Table 1. Compared to patients without diabetes, patients with diabetes were older (60±10 vs 56±12 years, p<0.001), more likely to have a history of hypertension (76 vs 52%, p<0.001), coronary artery disease (72 vs 52%, p<0.001) and had worse NYHA Functional Class (median Class II vs I, p=0.04). Patients with diabetes had higher creatinine (96 [81–118] vs 91 [77–105] μmol/L, p=0.02) and elevated cardiac biomarkers: NT-proBNP (1091 [326–2272] vs 579 [232–1136] pg/mL, p<0.001), hs-TnT (27 [14–41] versus 15 [9–22] ng/L, p<0.001), GDF-15 (2412 [1603–3331] versus 1039 [753–1470] pg/mL, p<0.001), sST2 (28 [23–38] versus 26 [21–32] ng/mL, p=0.038) and Gal-3 (10.0 [8.0-12.0] versus 8.0 [7.0-10.0] ng/mL, p<0.001). There was no difference in LV ejection fraction and GLS between the groups. The diabetes group had a higher prevalence of LGE (76 vs 60%, p=0.002, driven by higher rates of ischaemic LGE (51 vs 33%, p=0.003). Prevalence of non-ischaemic LGE did not differ between the groups. The diabetes group had higher native T1 (1285±42 vs 1269±42 ms, p<0.001) and ECV (30.5±3.5 vs 28.8±4.1%, p<0.001). After adjustment for potential confounders, including age, sex, hypertension and coronary artery disease, diabetes remained independently associated with the presence of LGE and increased ECV (p<0.005 for both analyses).

**Table 1.** Baseline clinical characteristics of the cohort according to presence or absence of diabetes.

|  | No Diabetes Mellitus<br>(n=156) | Diabetes Mellitus<br>(n=159) | p Value |
| --- | --- | --- | --- |
| **Clinical** |  |  |  |
| Age, years | 56 ± 12 | 60 ± 10 | <0.001 |
| Male sex, n (%) | 129 (83) | 126 (79) | 0.436 |
| Heart rate, beats/min | 71 [63-81] | 75 [68-83] | 0.020 |
| Systolic BP, mmHg | 124 [111-138] | 125 [113-143] | 0.240 |
| Diastolic BP, mmHg | 73 [66-82] | 70 [61-80] | 0.008 |
| Body mass index, kg/m<sup>2</sup> | 26 [23-30] | 27 [23-31] | 0.412 |
| Co-morbidities, n (%) |  |  |  |
| Hypertension | 80 (52) | 120 (76) | <0.001 |
| Coronary artery disease | 78 (52) | 112 (72) | <0.001 |
| Atrial Fibrillation | 22 (14) | 29 (18) | 0.331 |
| Medications at baseline, n (%) |  |  |  |
| Beta Blocker | 135 (87) | 146 (92) | 0.436 |
| ACEi or ARB | 129 (83) | 140 (88) | 0.178 |
| Mineralocorticoid antagonist | 98 (63) | 102 (64) | 0.806 |
| Diuretic | 116 (74) | 129 (81) | 0.148 |
| Statin | 120 (77) | 146 (92) | <0.001 |
| NYHA Functional Class, n (%) |  |  | 0.041 |
| Class I | 91 (58) | 75 (47) |  |
| Class II | 51 (33) | 74 (47) |  |
| Class III | 14 (9) | 10 (6) |  |
| MLWHF Score | 12 [0-30] | 14 [3-29] | 0.299 |
| Haematocrit, (%) | 43 ± 5 | 41 ± 6 | <0.001 |
| Creatinine, µmol/L | 91 [77-105] | 96 [81-118] | 0.016 |
| **Plasma Biomarkers** |  |  |  |
| NT-proBNP, pg/mL | 579 [232-1136] | 1091 [326-2272] | <0.001 |
| hs-TnT, ng/L | 15 [9-22] | 27 [14-41] | <0.001 |
| GDF-15, pg/mL | 1039 [753-1470] | 2412 [1603-3331] | <0.001 |
| sST2, ng/mL | 26 [21-32] | 28 [23-38] | 0.038 |
| Gal-3, ng/mL | 8.0 [7.0-10.0] | 10.0 [8.0-12.0] | <0.001 |
| **CMR Markers** |  |  |  |
| LVEDVi, mL/m<sup>2</sup> | 112 [83-142] | 99 [81-129] | 0.029 |
| LVESVi, mL/m<sup>2</sup> | 71 [43-105] | 65 [40-94] | 0.190 |
| SVi, mL/m<sup>2</sup> | 39 [32-44] | 35 [29-42] | 0.005 |
| LVMi, g/m<sup>2</sup> | 70 [59-89] | 67 [55-83] | 0.125 |
| LV ejection fraction, % | 36 [25-49] | 36 [26-49] | 0.947 |
| GLS, % | -10.1 ± 4.1 | -9.8 ± 3.7 | 0.443 |
| LAVi, mL/m<sup>2</sup> | 48 [38-68] | 47 [33-65] | 0.285 |
| LGE Type |  |  | 0.003 |
| Nil, n (%) | 63 (40) | 38 (24) |  |
| Non-ischaemic, n (%) | 38 (24) | 34 (21) |  |
| Ischaemic, n (%) | 52 (33) | 81 (51) |  |
| Both, n (%) | 3 (2) | 6 (4) |  |
| Native T1, ms | 1269 ± 42 | 1285 ± 42 | <0.001 |
| ECV, % | 28.8 ± 4.1 | 30.5 ± 3.5 | <0.001 |

Values are given as median [interquartile range], mean ± SD or number (percentage).
**Abbreviations:**; ACEi: Angiotensin-converting enzyme inhibitor; ARB: Angiotensin II receptor blocker; NYHA: New York Heart Association; MLWHF: Minnesota Living with Heart Failure; NT-proBNP: N-terminal pro-brain natriuretic peptide; hs-TnT: high-sensitivity troponin T; GDF-15: growth differentiation factor 15; sST2: soluble ST2; Gal-3: galectin 3; LVEDVi: Left ventricular end-diastolic volume indexed to body surface area; LVESVi: Left ventricular end-systolic volume index; SVi: Stroke volume index; LVMi: Left ventricular mass index; GLS: Global longitudinal strain; LAVi: Left atrial end-systolic volume index; LGE: Late gadolinium enhancement; ECV: Extracellular volume.

### Predictors of Primary Outcome in All Patients with Heart Failure

Over a median follow-up of 23 [18–24] months, 74 patients experienced the primary outcome (52 hospitalisations for HF, 22 all-cause deaths). In univariable Cox regression analyses for the entire cohort, clinical markers associated with the primary outcome included diabetes, NYHA functional class and systolic blood pressure. Circulating biomarkers associated with the primary outcome included NT-proBNP, hs-TnT, GDF-15, sST2 and Gal-3. CMR markers of adverse remodelling (LV mass index and end diastolic volume index), contractile function (LV ejection fraction and GLS) and myocardial fibrosis (presence of LGE, native T1 and ECV) were predictors of worse outcomes. Diabetes, NT-proBNP and GLS remained independently associated with outcomes in the multivariable analysis (Table 2 and Figure 2).

**Figure 2.**
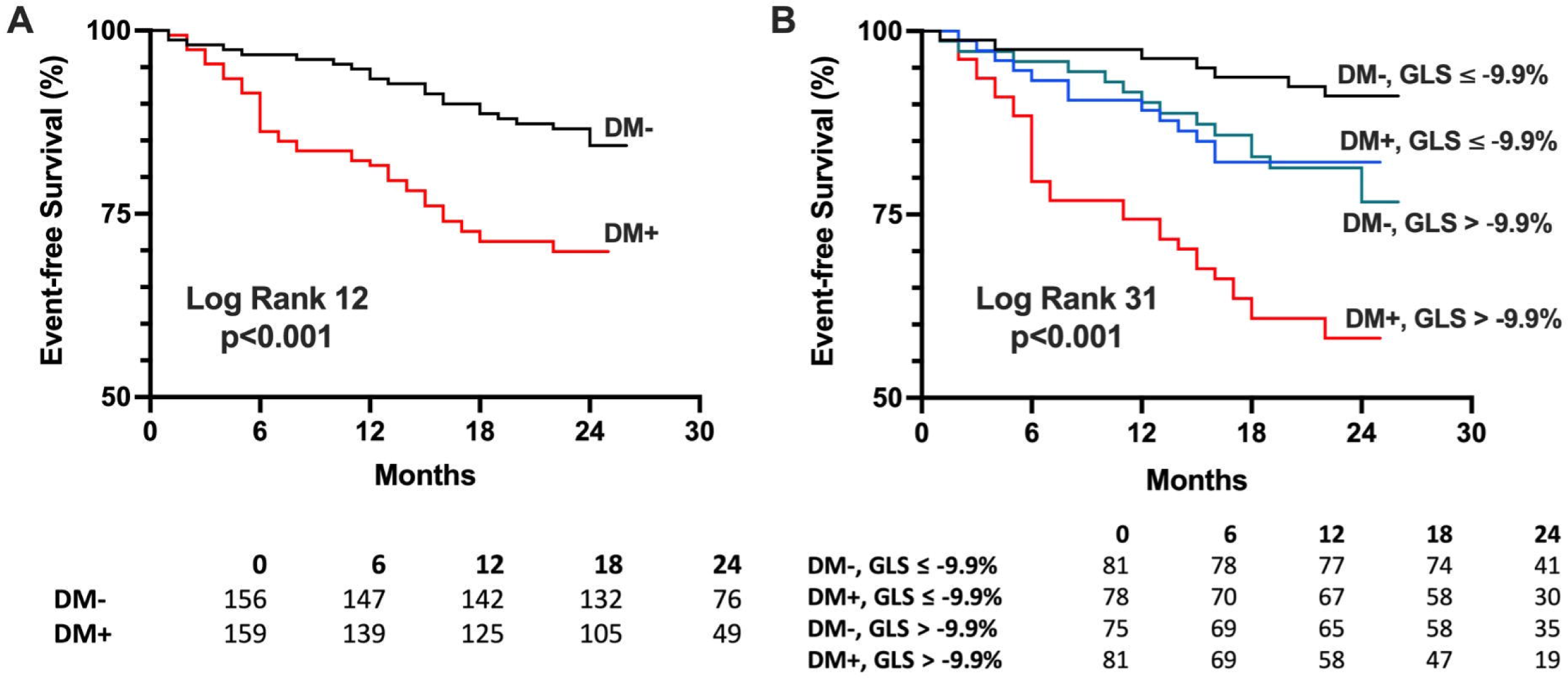
Event-free survival curves showing: (A) Adverse prognosis in patients with heart failure and diabetes; (B) Patients with diabetes and GLS worse than cohort median (-9.9%) had worst prognosis. Patients with either diabetes or GLS worse than median had similar outcomes. Abbreviations: DM: diabetes mellitus; GLS: global longitudinal strain

**Table 2.**
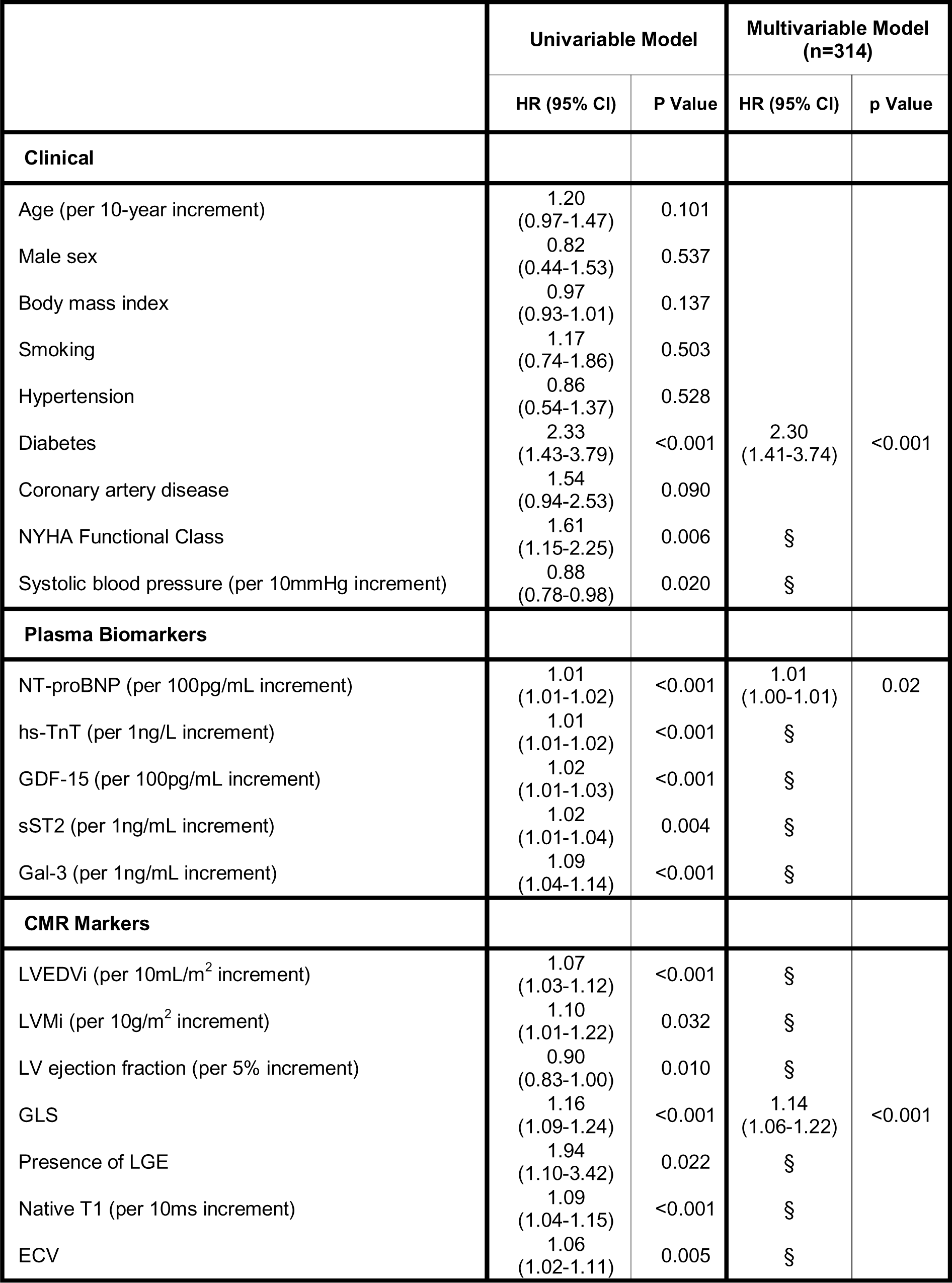
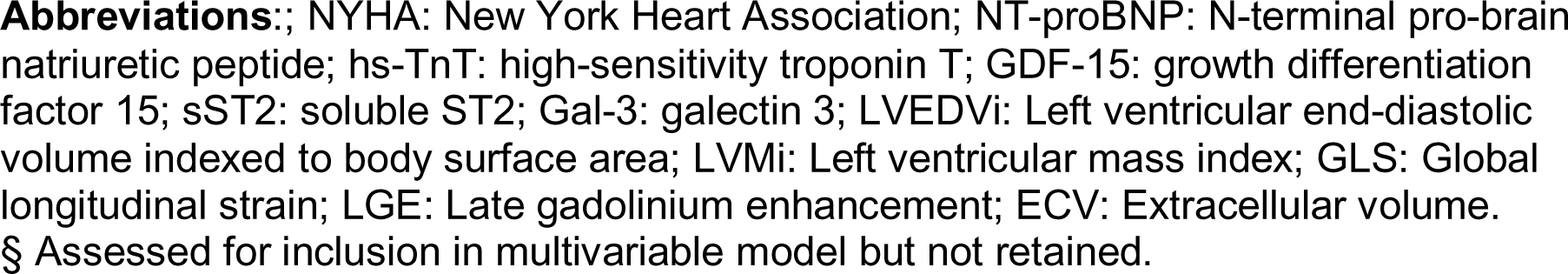
Univariable and multivariable Cox regression models (stepwise selection) in the entire cohort for the primary outcome of all-cause mortality or heart failure hospitalisation.

### Predictors of Primary Outcome in Patients with Heart Failure and Diabetes

In the diabetes group, 50 patients experienced the primary outcome (35 hospitalisations for HF, 15 all-cause deaths). In univariable analyses, systolic blood pressure, NT-proBNP, hs-TnT, GDF-15, sST2, LV ejection fraction, GLS and native T1 were associated with the primary outcome (Table 3). GLS and sST2 remained associated with outcomes in the multivariable analysis. Patients with diabetes and worse than median GLS (GLS > -9.9%) had the worst prognosis (log-rank p<0.001, Figure 3). Of note, patients with diabetes and GLS better than median had similar outcomes to patients without diabetes and GLS worse than median (Figure 2B; p=0.70).

**Figure 3.**
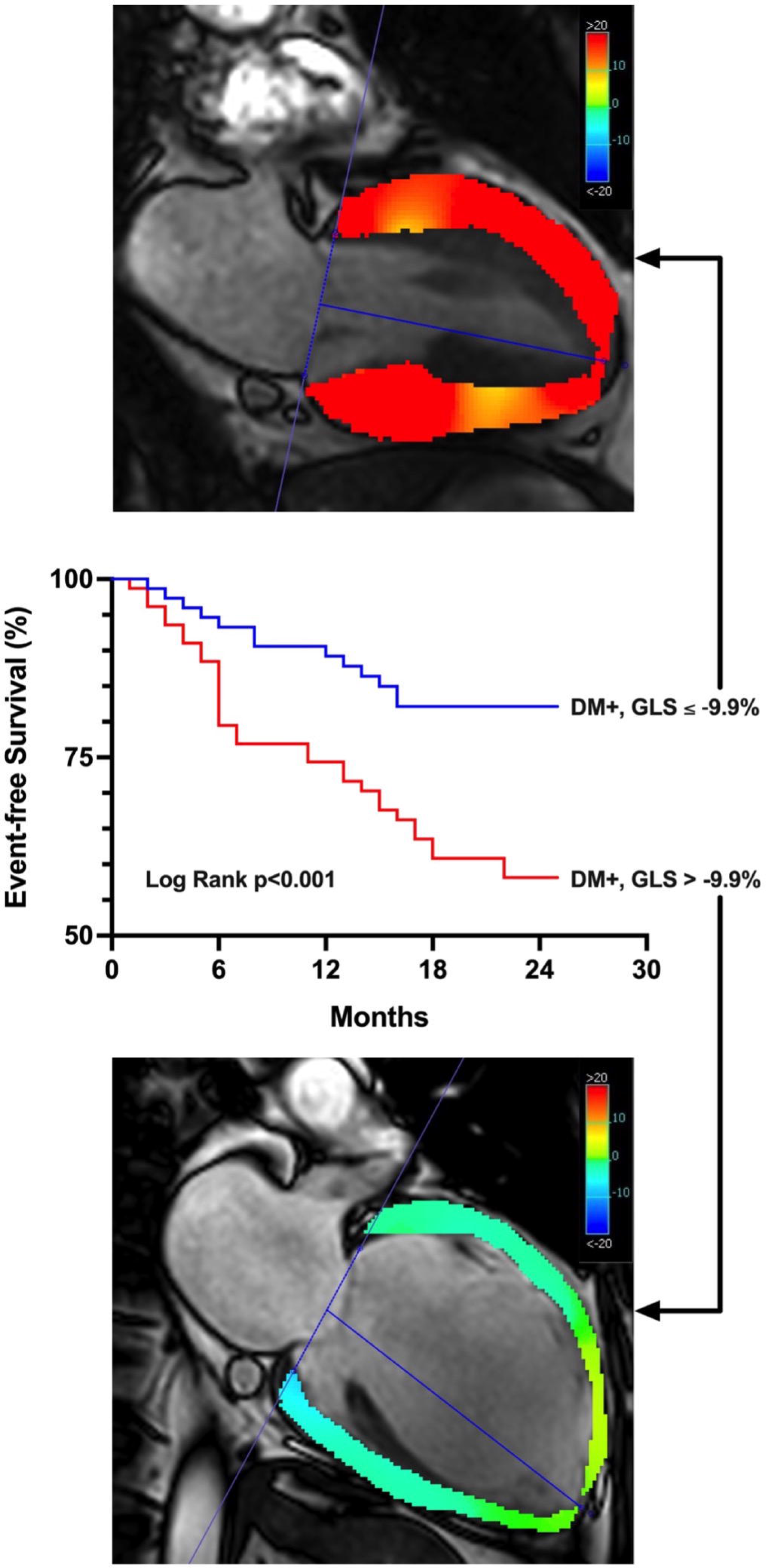
Example of GLS measurement in apical 2-chamber view. **Top panel** shows a patient with heart failure, diabetes and GLS ≤ -9.9%. **Bottom panel** shows a patient with heart failure, diabetes and GLS > -9.9%. **Centre panel:** event-free survival curves showing adverse prognosis in patients with heart failure, diabetes and GLS worse than cohort median (-9.9%). Abbreviations: DM: diabetes mellitus; GLS: global longitudinal strain.

**Table 3.** Univariable and multivariable Cox regression models (stepwise selection) in the diabetes group for the primary outcome of all-cause mortality or heart failure hospitalisation.

|  | Univariable Model |  | Multivariable Model (n=158) |  |
| --- | --- | --- | --- | --- |
|  | HR (95% CI) | P Value | HR (95% CI) | p Value |
| **Clinical** |  |  |  |  |
| Age (per 10-year increment) | 1.21<br>(0.90-1.60) | 0.202 |  |  |
| Male sex | 0.85<br>(0.41-1.75) | 0.665 |  |  |
| Body mass index | 0.99<br>(0.94-1.05) | 0.827 |  |  |
| Hypertension | 0.94<br>(0.50-1.76) | 0.838 |  |  |
| Diabetes Type | 0.88<br>(0.28-2.84) | 0.836 |  |  |
| Smoking | 0.80<br>(0.46-1.40) | 0.436 |  |  |
| Coronary artery disease | 1.41<br>(0.72-2.75) | 0.314 |  |  |
| NYHA Functional Class | 1.34<br>(0.86-2.08) | 0.200 |  |  |
| Systolic blood pressure (per 10mmHg increment) | 0.86<br>(0.75-0.98) | 0.028 | § |  |
| **Plasma Biomarkers** |  |  |  |  |
| NT-proBNP (per 100pg/mL increment) | 1.01<br>(1.00-1.02) | 0.007 | § |  |
| hs-TnT (per 1ng/L increment) | 1.01<br>(1.00-1.02) | 0.016 | § |  |
| GDF-15 (per 100pg/mL increment) | 1.02<br>(1.01-1.03) | 0.003 | § |  |
| sST2 (per 1ng/mL increment) | 1.03<br>(1.01-1.06) | 0.003 | 1.03<br>(1.00-1.06) | 0.02 |
| Gal-3 (per 1ng/mL increment) | 1.06<br>(1.00-1.12) | 0.061 |  |  |
| **CMR Markers** |  |  |  |  |
| LVEDVi (per 10mL/m<sup>2</sup> increment) | 1.07<br>(0.99-1.15) | 0.076 |  |  |
| LVMi (per 10g/m<sup>2</sup> increment) | 1.01<br>(0.88-1.15) | 0.928 |  |  |
| LV ejection fraction (per 5% increment) | 0.89<br>(0.81-0.99) | 0.031 | § |  |
| GLS | 1.15<br>(1.06-1.25) | 0.001 | 1.12<br>(1.03-1.21) | 0.01 |
| Presence of LGE | 1.42<br>(0.69-2.92) | 0.342 |  |  |
| Native T1 (per 10ms increment) | 1.07<br>(1.00-1.14) | 0.04 | § |  |
| ECV | 1.06<br>(0.98-1.15) | 0.139 |  |  |

**Abbreviations:**; NYHA: New York Heart Association; NT-proBNP: N-terminal pro-brain natriuretic peptide; hs-TnT: high-sensitivity troponin T; GDF-15: growth differentiation factor 15; sST2: soluble ST2; Gal-3: galectin 3; LVEDVi: Left ventricular end-diastolic volume indexed to body surface area; LVMI: Left ventricular mass index; GLS: Global longitudinal strain; LGE: Late gadolinium enhancement; ECV: Extracellular volume. § Assessed for inclusion in multivariable model but not retained.

In a multivariable model including clinical markers and plasma biomarkers, only sST2 remained independently associated with the primary outcome, demonstrating stronger prognostic associations than NT-proBNP (Table 4).

**Table 4.** Multivariable Cox regression model (Clinical and Plasma biomarkers) in the diabetes group for the primary outcome of all-cause mortality or heart failure hospitalisation.

|  | Multivariable Model<br>(n=158) |  |
| --- | --- | --- |
|  | HR (95% CI) | p Value |
| **Clinical** |  |  |
| Age (per 10-year increment) |  |  |
| Male sex |  |  |
| Body mass index |  |  |
| Hypertension |  |  |
| Diabetes Type |  |  |
| Smoking |  |  |
| Coronary artery disease |  |  |
| NYHA Functional Class |  |  |
| Systolic blood pressure (per 10mmHg increment) | § |  |
| **Plasma Biomarkers** |  |  |
| NT-proBNP (per 100pg/mL increment) | § |  |
| high-sensitivity troponin T | § |  |
| GDF-15 (per 100pg/mL increment) | § |  |
| sST2 | 1.04<br>(1.02-1.07) | 0.001 |
| Gal-3 |  |  |

**Abbreviations:**; NYHA: New York Heart Association; NT-proBNP: N-terminal pro-brain natriuretic peptide; hs-TnT: high-sensitivity troponin T; GDF-15: growth differentiation factor 15; sST2: soluble ST2; Gal-3: galectin 3.
§ Assessed for inclusion in multivariable model but not retained.

The diabetes group was also stratified into four groups based on GLS (above or below diabetes group median of -9.7%) and plasma biomarker (above or below median for NT-proBNP, hs-TnT, GDF-15, sST2 and Gal-3). Amongst patients with above median GLS, NT-proBNP and GDF-15 showed additional prognostic value (Supplemental Figure 1). On the other hand, the combination of below median GLS and sST2 demonstrated particularly adverse prognosis (Supplemental Figure 2).

## Discussion

The main finding of this study is that GLS is a powerful independent predictor of adverse outcomes in patients with HF and diabetes, providing incremental prognostic information beyond several circulating plasma biomarkers and CMR markers of cardiac remodelling, inflammation and fibrosis. In the diabetes group, worse GLS (> median -9.9%) was associated with adverse prognosis. GLS is known to have prognostic value in HF, regardless of whether the cause is ischemic or non-ischemic and whether the EF is reduced or preserved [14, 15]. In this study, we have extended those findings to the diabetes subgroup, who are known to have a higher risk of adverse cardiovascular outcomes. Our findings are in agreement with a prior study which showed that speckle tracking echocardiography (STE) GLS has prognostic value in diabetes patients with dilated cardiomyopathy [28]. The results from the current study raise the possibility of GLS-guided risk stratification and management in patients with HF and diabetes. Indeed, there is emerging evidence of improvement in GLS with anti-diabetic medications which may enable this in future [28–30].

GLS by CMR is well-suited for routine clinical use. The technique relies on software packages which track the endocardial and epicardial borders, and reference values are specific for the software used for analysis. STE has a number of limitations, including dependence on high quality 2D images, and results are often affected by foreshortening, dropout of the apical and anterolateral segments on apical views, and/or suboptimal acoustic windows [15, 31, 32]. GLS by CMR overcomes these limitations and can be measured from routinely acquired bright-blood, steady-state free precession long-axis cine imaging. Disadvantages of GLS by CMR include its susceptibility to through-plane motion artefacts, limited temporal resolution for quantification of strain rate, and limitations in patients with contraindications to CMR [15]. Furthermore, GLS by CMR values may be affected by inter-vendor differences in algorithms, similar to STE, which has resulted in difficulties establishing reference values [32]. Nonetheless, GLS has shown close correlation with STE and has become an important component of the CMR examination alongside volumetric analysis and tissue characterisation in the assessment of HF [33].

In this study, sST2 demonstrated incremental prognostic value over other biomarkers including NT-proBNP and hs-TnT in HF patients with diabetes. sST2 is produced by cardiomyocytes and cardiac fibroblasts in response to myocardial stress, inflammation or injury [34–36]. Other sources of sST2 include endothelial cells of the aorta and coronary arteries as well as certain immune cells [34]. It acts as a decoy receptor for interleukin-33, attenuating its cardioprotective biological effects which include blunting myocardial hypertrophy and fibrosis, and inhibiting cardiomyocyte apoptosis [18, 34, 37]. We have confirmed that sST2 levels are higher in HF patients with diabetes [38, 39]. Furthermore, sST2 has known prognostic value in acute and chronic HF, independent of plasma natriuretic peptides [40–42]. We have extended these findings to the subgroup of HF patients with diabetes. This result provides further strength to the possibility that the prognostic value of sST2 in heart failure may result from its association with metabolic perturbances that are also commonly found in diabetes [43]. Our findings therefore suggest a possible role for sST2 in risk stratification amongst HF patients with diabetes and in monitoring response to treatment [44–49].

Diabetes was a strong independent predictor of the composite outcome of hospitalisation for HF or all-cause mortality in this HF cohort. This is in agreement with prior studies which have consistently shown worse cardiovascular outcomes in HF patients with diabetes, regardless of HF etiology and whether the ejection fraction is reduced or preserved [8, 50, 51]. Furthermore, Kaplan-Meier survival analysis showed similar event rates in the diabetes group with above median GLS (≤ -9.9%) compared to those without diabetes and below median GLS. These findings are similar to data from the Candesartan in Heart failure: Assessment of Reduction in Mortality and morbidity (CHARM) study, which demonstrated that patients with HF, preserved ejection fraction (EF> 40%), and diabetes had a greater rate of HF hospitalisation than those with lower ejection fraction (EF≤40%) and no diabetes [8]. These findings highlight the urgent need for therapeutic advances in patients with HF and concomitant diabetes.

In this study, HF patients with diabetes had worse CMR markers of myocardial injury, inflammation, and fibrosis. The association between diabetes and elevated ECV remained after adjusting for potential confounders that differed between the diabetic and non-diabetic groups including age, hypertension, coronary artery disease and presence of LGE. Furthermore, diabetes remained associated with the presence of LGE in logistic regression models, even accounting for age, sex, hypertension and coronary artery disease. These findings are in agreement with prior studies, including a recent meta-analysis, which showed an association between diabetes and a higher degree of myocardial fibrosis as assessed by histology as well as ECV by CMR [11].

Both focal and diffuse myocardial fibrotic processes are known to occur in patients with diabetes independently of co-morbid conditions, including coronary atherosclerosis and hypertension. Diffuse interstitial and perivascular fibrosis are structural hallmarks of diabetic cardiomyopathy, but focal replacement fibrosis can also be seen, even in the absence of coronary artery disease [52]. Several mechanisms may explain the fibrosis burden in diabetes. Hyperglycaemia is thought to upregulate the expression of profibrotic factors such as transforming growth factor beta 1 and down-regulate the activity of the matrix metalloproteinases [53]. Hyperglycaemia is also known to promote the formation of advanced glycation end products (AGEs) which can cross-link collagen in the interstitium, increasing their resistance to degradation. AGEs can result in generation of reactive oxygen species and oxidative stress which further promotes a pro-fibrotic state [11]. Pro-inflammatory cytokines and chemokines, as well as increased renin-angiotensin-aldosterone system activity in diabetes have also been implicated in the development of myocardial fibrosis.

Native T1 values were higher in the diabetes group and may reflect interstitial expansion due to myocardial fibrosis as well as myocardial oedema affecting the cellular and interstitial compartments [54]. There is some disagreement in the literature regarding whether native T1 values are increased in diabetes. Several studies have shown an association between diabetes and increased T1 [55–58]. Although a positive association was found between diabetes and native T1 in a recent meta-analysis, the result was not statistically significant [11]. The authors of that study suggested that the lack of statistical significance may have resulted from limited sample size of the included studies. Indeed, the diabetes cohort in the present study was larger than any of those in the included studies. Furthermore, T1 values are known to be dependent on a variety of factors, including field strength, pulse sequence, and region of measurement within the myocardium. In the present study, T1 maps were obtained at 3T using the same MOLLI sequence for all patients, and with experienced observers performing standardized analyses that may improve reproducibility and eliminate technical cofounders [59, 60]. Whilst it does appear that diabetes is associated with higher T1, larger studies controlling for the variability in T1 are required to more conclusively answer this question.

### Limitations

One limitation of this study is the absence of measures of glycaemic control, which are known to be prognostic in patients with HF and diabetes [61]. However, STE GLS is known to have prognostic value independent of glycaemic control in diabetes cohorts with preserved and reduced ejection fraction, and therefore this is unlikely to have altered the prognostic associations for GLS by CMR [16, 28]. Our HF cohort included patients with both ischaemic and non-ischaemic etiologies, as well as both preserved and reduced ejection fraction. Unfortunately, the study was not powered for analyses of these subgroups. Documented history of coronary artery disease did not associate with outcomes in the univariable analysis and therefore etiology of HF is unlikely to have affected the results. Finally, this was a single centre study using feature tracking software from a single vendor, limiting generalizability of the results.

## Conclusions

Patients with HF and diabetes had worse CMR and plasma markers of injury, inflammation, and fibrosis, and an adverse prognosis. sST2 showed incremental prognostic value beyond NT-proBNP in HF patients with diabetes. GLS is an important and independent prognostic marker in this group. Future studies should explore whether GLS-guided risk stratification and management can improve outcomes in this group of patients.

## Data Availability

The data that support the findings of this study are included
in this article or available from the corresponding author upon
reasonable request.

## Abbreviations

HF: heart failure
GLS: global longitudinal strain
CMR: cardiovascular magnetic resonance
LGE: late gadolinium enhancement
ECV: extraceullar volume
NT-proBNP: N-terminal pro B-type natriuretic peptide
hs-TnT: high-sensitivity troponin T
GDF-15: growth differentiation factor 15
sST2: soluble ST2
Gal-3: galectin 3
left ventricle: LV

## Declarations

### Ethics approval and consent to participate

Ethics approval was obtained from the local Centralized Institutional Review Board in Singapore, and all participants provided written informed consent. The study was conducted in accordance with the principles of the Declaration of Helsinki.

### Consent for publication

Consent for the publication of images was given.

### Availability of data and materials

The datasets generated and analysed for the current study are not publicly available. Please contact the corresponding author for data requests.

### Authors’ contributions

CSPL and AMR designed the study. PJC, FE, RF and CWLC made substantial contributions to the study design. NRI, SPC, OWL, JPCC, JAB and TTL collected the data. NRI analysed the data. NRI, MU and CWLC interpreted the data and wrote the manuscript. CC and AMR made substantial revisions to the manuscript. All authors read and approved the final manuscript.

## Acknowledgements

The contribution of all the site investigators and clinical co-ordinators is acknowledged.

## Supplemental Figures

**Supplemental Figure 1. Event-free survival curves in patients with diabetes and above median GLS (-9.7%) stratified by:** (A) NT-proBNP; (B) hs-TnT; (C) GDF-15; (D) sST2 and (E) Gal-3. NT-proBNP and GDF-15 demonstrated additional prognostic value in this group. Abbreviations: GLS: global longitudinal strain; NT-proBNP: N-terminal pro-brain natriuretic peptide; hs-TnT: high-sensitivity troponin T; GDF-15: growth differentiation factor 15; sST2: soluble ST2; Gal-3: galectin 3.

**Supplemental Figure 2. Event-free survival curves in patients with diabetes and below median GLS (-9.7%) stratified by:** (A) NT-proBNP; (B) hs-TnT; (C) GDF-15; (D) sST2 and (E) Gal-3. sST2 demonstrated additional prognostic value in this group. Abbreviations: GLS: global longitudinal strain; NT-proBNP: N-terminal pro-brain natriuretic peptide; hs-TnT: high-sensitivity troponin T; GDF-15: growth differentiation factor 15; sST2: soluble ST2; Gal-3: galectin 3.

