## Supplementary figures and images for "Global longitudinal strain and plasma biomarkers for prognosis in heart failure complicated by diabetes"

### Supplemental Figure 1

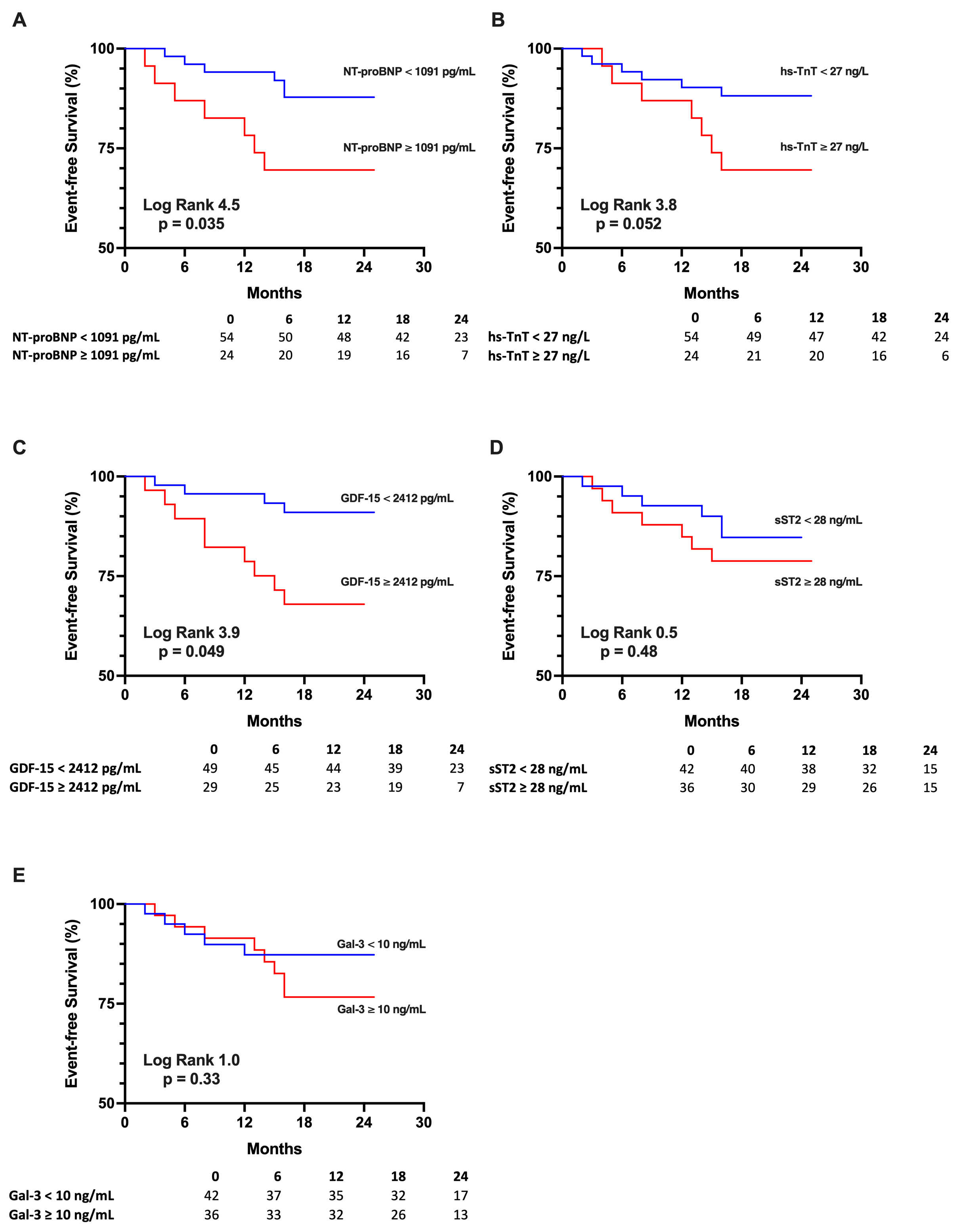

### Supplemental Figure 2

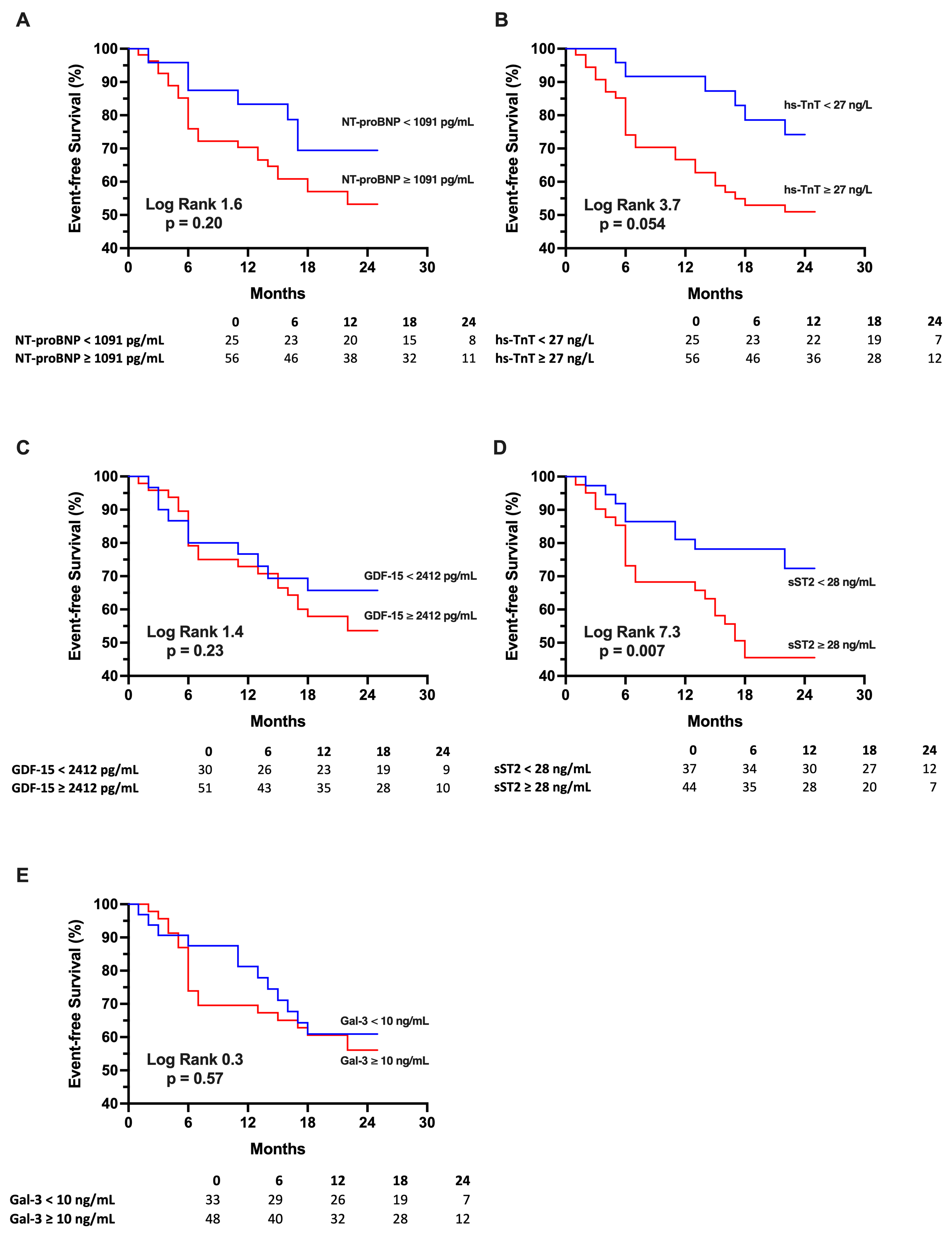
